# Plasma and CSF neurofilament light chain distinguish neurodegenerative from primary psychiatric conditions in a clinical setting

**DOI:** 10.1101/2024.08.11.24311847

**Authors:** Dhamidhu Eratne, Matthew JY Kang, Courtney Lewis, Christa Dang, Charles B Malpas, Michael Keem, Jasleen Grewal, Vladimir Marinov, Amy Coe, Cath Kaylor-Hughes, Thomas Borchard, Chhoa Keng-Hong, Alexandra Waxmann, Burcu Saglam, Tomas Kalincik, Richard Kanaan, Wendy Kelso, Andrew Evans, Sarah Farrand, Samantha Loi, Mark Walterfang, Christiane Stehmann, Qiao-Xin Li, Steven Collins, Colin L Masters, Alexander F Santillo, Henrik Zetterberg, Kaj Blennow, Samuel F Berkovic, Dennis Velakoulis, The MiND Study Group

## Abstract

**INTRODUCTION:** Many patients with neurodegenerative disorders (ND) face diagnostic delay and misdiagnosis. We investigated blood and cerebrospinal fluid (CSF) neurofilament light chain (NfL) to distinguish ND from primary psychiatric disorders (ND), a common challenge in clinical settings.

**METHODS:** Plasma and CSF NfL levels were measured and compared between groups, adjusting for age, sex, weight.

**RESULTS:** 337 participants included: 136 ND, 77 PPD, 124 Controls. Plasma NfL was 2.5 fold elevated in ND compared to PPD and had strong diagnostic performance (area under the curve, AUC 0.86, 81%/85% specificity/sensitivity) that was comparable to CSF NfL (2 fold elevated, AUC 0.89, 95%/71% specificity/sensitivity). Diagnostic performance was especially strong in younger people (40-<60years). Additional findings were cut-offs optimised for sensitivity and specificity, and issues important for future clinical translation

**CONCLUSIONS:** This study adds important evidence for a simple blood-based biomarker to assist as a screening test for neurodegeneration and distinction from PPD, in clinical settings.

## INTRODUCTION

Despite major improvements in clinical assessment, many patients with neurodegenerative disorders (ND) still face significant barriers and challenges to timely, accurate diagnosis; delays often last several years even with gold standard assessments [1]. These challenges are worse for younger people (onset of symptoms <60-65 years of age), where a wider range of ND and less typical presentations of ND are more common, and substantial overlap in symptoms with psychiatric disorders exists [1–3]. Methods to distinguish ND from primary psychiatric disorders (PPD) are a major unmet need.

One of the most well-established biomarkers for neuronal injury, neurofilament light chain protein (NfL), has shown great promise in distinguishing broad causes of ND from PPD and non-neurodegenerative disorders. We have previously demonstrated the strong diagnostic utility of cerebrospinal fluid (CSF) NfL [3–7]; however, a blood-based biomarker could be a less invasive, more easily accessible option to improve diagnosis, which is critical for improved outcomes for patients, families, healthcare systems, and clinical trials.

Research investigating blood NfL in distinguishing diverse ND directly from diverse PPD in clinical settings, especially in younger populations, has been limited. Most studies have focused on comparing NfL levels between different NDs and controls [8–13] Some studies have investigated CSF and blood NfL for broad ND diagnosis in clinical settings, finding elevated concentrations of NfL in ND, diagnostic utility and/or increasing diagnostic certainty [14–20]. The latter studies have primarily been in older people in memory clinic settings, and most have not specifically been compared to diverse PPD. Studies that have included PPD have either been small (e.g., n=17) [20], or grouped PPD with healthy controls or “non-neurodegenerative disorder” or similar categorisations, and did not specifically compare ND to PPD [14], despite some evidence that plasma NfL may be mildly elevated in PPD compared to healthy controls [21,22]. While blood NfL has shown strong diagnostic performance in distinguishing PPD from specific ND subtypes, such as behavioural variant frontotemporal dementia (bvFTD) [8,9,21–24], broader comparative studies across diverse ND and PPD populations are needed to fully realise its clinical utility, and for real-world clinical translation. Finally, there is also increasing recognition of the importance of further research in clinical settings, including important covariates that can influence individual levels and reference ranges and interpretations, such as age, analytical platform and technological factors, for standardisation and proper clinical translation [25–29].

This study aims to address the significant gaps in research by comparing diverse ND and PPD reflective of real-world practice, with a focus on younger populations where the diagnostic overlap between ND and PPD is particularly challenging, aiming to provide a more nuanced understanding of NfL’s diagnostic utility in a clinical setting. The primary aim of this study was to investigate differences in blood and CSF NfL concentrations between ND and PPD seen in a clinical neuropsychiatry service, and their diagnostic performance in differentiating between the two diagnostic groups (Aim 1). We also aimed to investigate the accuracy and utility of age-based cut-off levels/concentrations between younger and older patients, and cut-offs optimising sensitivity and specificity (Aim 2). For issues related to clinical translation, we also aimed to perform exploratory analyses to compare accuracy of different classification systems, including our previously described age-adjusted percentile and z-score models [22], and previously described reference ranges (Aim 3).

## METHODS

### Study cohorts

This study included participants prospectively recruited between June 2019 and April 2023, who had provided a blood sample for NfL analysis. A subset of patients had CSF collected for clinical purposes, with remnant samples available for NfL analysis. The patient cohorts were people referred for diagnostic assessment and management of possible ND to the Neuropsychiatry Centre at The Royal Melbourne Hospital, a quaternary service receiving referrals for diagnostically complex cases from primary care and other specialist services within Australia. Patients received comprehensive multidisciplinary assessments and multimodal investigations, including CSF analysis, with gold standard consensus diagnosis based on established diagnostic criteria, as previously described in detail [3,5]. Control participants were people recruited from the community, with no symptoms or diagnoses of neurological or neurodegenerative disorders, no active psychiatric symptoms or conditions.

This study included 38 patients (26 ND, 12 PPD) from our previous CSF study [3]. The remaining patients (n=175) controls (n=124), and data in this study, (including all blood NfL data), have not been described previously. Diagnostic group categorisation was determined based on most recent diagnosis at longitudinal follow up, blinded to NfL levels, as previously described [3,5].

CSF and EDTA plasma samples were stored at –80C. Plasma NfL was measured using NF-Light kits on a Quanterix Single molecule array (Simoa) HD-X analyzer, according to the manufacturer’s instructions (Quanterix Corporation, Billerica, MA, USA). CSF NfL was measured using a commercial enzyme-linked immunosorbent assay (ELISA; NF-light; UmanDiagnostics, Sweden).

### Statistical analyses

Statistical analyses were performed using R version 4.3.2 (2023-10-31). As several biomarker distributions were non-Gaussian even when log-transformed, biomarker levels in different groups were compared using standardised bootstrapped quantile regression, with age and sex as additional covariates. To examine statistical effects, standardised quantile regression coefficients (ß) were interpreted along with 95% confidence intervals (CIs). ROC curve analyses were performed to investigate diagnostic utility between different combinations of groups. Bootstrapped DeLong test was used to compare ROC curves. Optimal cut-offs were selected based on Youden’s J, and alternative cut-offs for screening based on optimised specificity and sensitivity. Additional diagnostic test parameters were computed: positive and negative likelihood ratios, positive and negative predictive values, overall accuracy, and diagnostic odds ratio (DOR). DOR is a single indicator of test performance that combines the sensitivity and specificity of a diagnostic test, reflecting the odds of a positive test result in patients with ND relative to the odds of a positive test result in those without. A higher DOR indicates better discriminatory test performance, with a DOR of 1 indicating that the test does not discriminate between patients with and without ND. Additional sensitivity analyses were performed: excluding extreme outliers and performing all quantile regressions with weight included as a covariate. As results were similar, the results excluding weight were presented to maximise the sample sizes for analyses and presented results (since not all participants had weight data).

This study, part of The Markers in Neuropsychiatric Disorders Study (The MiND Study, https://themindstudy.org), was approved by the Human Research Ethics Committee at Melbourne Health (2016.038, 2017.090, 2018.371, 2020.142).

## RESULTS

### Study cohort

The total study cohort included 337 participants: 136 with ND, 77 with PPD, and 124 Controls (Table 1). Controls were slightly older at 63.2 years compared to the other groups (ND 60.8 years, PPD 54.8 years), and had a greater proportion of females (71% compared to ND 43%, PPD 51%). 250 people had weight data. PPD had higher weight (84kg) compared to ND (75kg) and Controls (76kg).

**Table 1.**
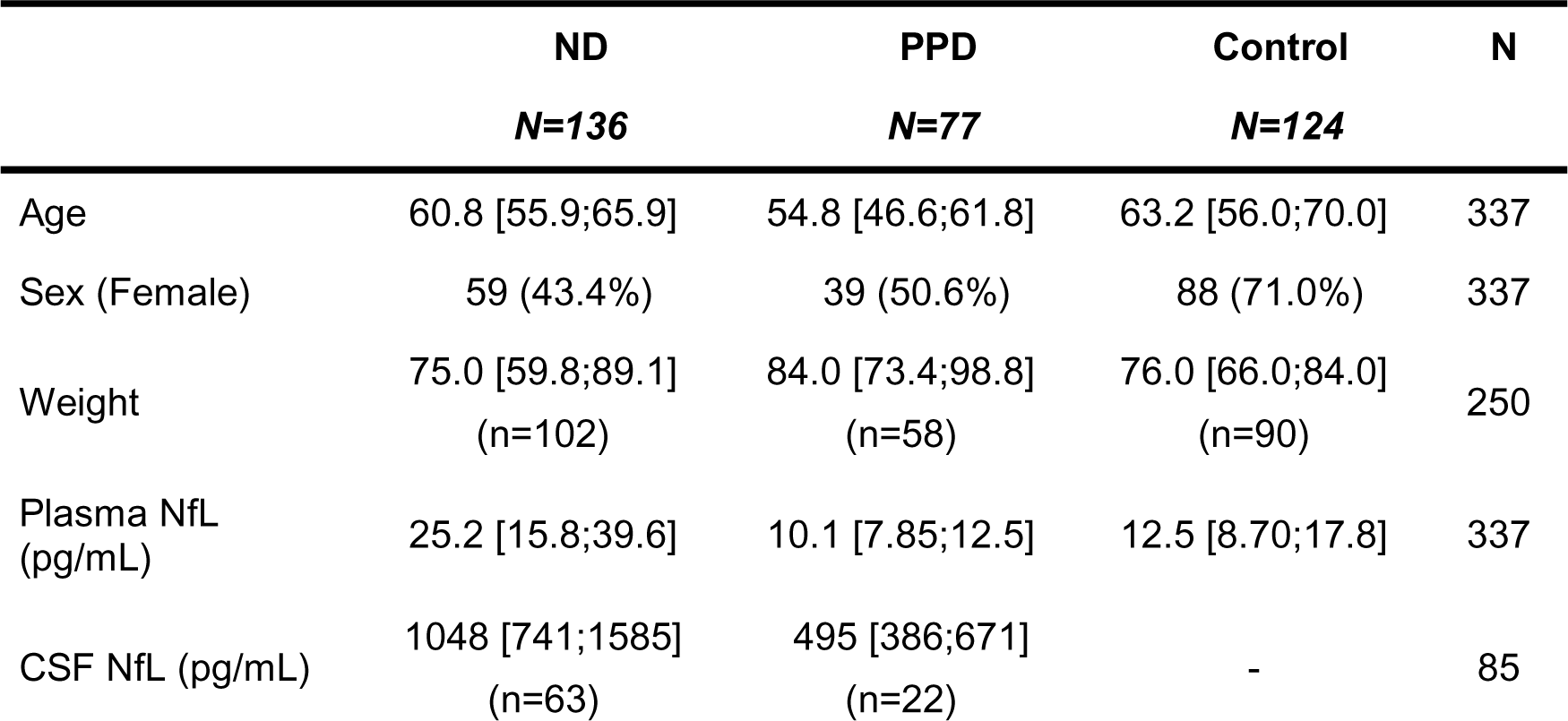
Study demographics and plasma and CSF neurofilament light (NfL) levels in neurodegenerative disorders (ND), primary psychiatric disorders (PPD), and Controls. Data are median [interquartile range] or n (%). CSF: cerebrospinal fluid; NfL: neurofilament light chain; ND: neurodegenerative disorder; PPD: primary psychiatric disorder

|  | **ND**<br>**N=136** | **PPD**<br>**N=77** | **Control**<br>**N=124** | **N** |
| --- | --- | --- | --- | --- |
| Age | 60.8 [55.9;65.9] | 54.8 [46.6;61.8] | 63.2 [56.0;70.0] | 337 |
| Sex (Female) | 59 (43.4%) | 39 (50.6%) | 88 (71.0%) | 337 |
| Weight | 75.0 [59.8;89.1]<br>(n=102) | 84.0 [73.4;98.8]<br>(n=58) | 76.0 [66.0;84.0]<br>(n=90) | 250 |
| Plasma NfL<br>(pg/mL) | 25.2 [15.8;39.6] | 10.1 [7.85;12.5] | 12.5 [8.70;17.8] | 337 |
| CSF NfL (pg/mL) | 1048 [741;1585]<br>(n=63) | 495 [386;671]<br>(n=22) | - | 85 |

All 337 people had plasma NfL. Both plasma and CSF NfL concentrations had been determined for 84 people (plasma+CSF group: 63 ND, 22 PPD), the remainder had only plasma NfL data available (plasma-only group: 252 people, ND 73, PPD 55, 124 Controls). Details of these subsets can be found in Supplementary Tables 1 and 2.

The ND group consisted of Alzheimer’s disease (AD, n=44), behavioural variant frontotemporal dementia (bvFTD, n=16), Lewy body dementia (DLB, n=7), dementia not-otherwise-specified (dementia NOS, n=9), Huntington’s disease (n=17), vascular dementia (n=6), mixed Alzheimer/vascular dementia (n=3), Creutzfeldt-Jakob disease (n=2), substance-induced dementia (n=2), and Other ND (n=31, which included autoimmune encephalitis, cerebral amyloid angiopathy, corticobasal syndrome, CNS vasculitis, Down syndrome, Fahr disease, metabolic disorders, Niemann-Pick Type C, Parkinson’s disease and cerebellar degenerative disorder). The PPD group consisted of major depressive disorder (MDD, n=23), schizophrenia spectrum disorders (n=17), functional neurological/cognitive disorders (n=8), bipolar affective disorder (BPAD, n=6), and Other PPD (n=21, including anxiety, obsessive compulsive, and post-traumatic stress disorders).

### Aim 1: Plasma and CSF NfL levels and diagnostic utility in neurodegenerative and primary psychiatric disorders

#### In subset of patients with both CSF and plasma samples, compared to subset with only plasma

We first separately analysed the subset of patients with both CSF and plasma levels (plasma+CSF), and the subset with only plasma levels (plasma-only), to determine comparability, and thus determine whether the total cohort could be analysed as a whole.

For the plasma+CSF group (n=85, 63 ND, 22 PPD), NfL levels were significantly elevated in ND compared to PPD, in CSF (median 1048pg/mL vs 495pg/mL, standardised quantile regression coefficient ß: 0.09, 95%CI [0.04, 0.53], p<0.001), and in plasma (median 24.4pg/mL. vs 10.3pg/mL, ß: 0.07, 95%CI [0.03, 0.54], p<0.001), Supplementary Table 1.

The plasma-only group (n=128, 73 ND, 55 PPD) had comparable results, with plasma NfL significantly elevated in ND compared to PPD (median 26.8pg/mL vs 10.1pg/mL, ß: 0.18, 95%CI [0.08, 0.70], p<0.001), Supplementary Table 2. We therefore analysed the entire cohort and present detailed results below, with a focus on plasma NfL.

#### In entire cohort

NfL levels were significantly elevated in ND compared to PPD, 2.5 fold in plasma (median 25.2pg/mL vs 10.1pg/mL; ß: 0.10 [0.05, 0.43], p<0.001), and 2 fold in CSF (median 1048pg/mL vs 495pg/mL, ß: 0.09, 95%CI [0.04, 0.53], p<0.001), Table 1 and Figure 1.

**Figure 1.**
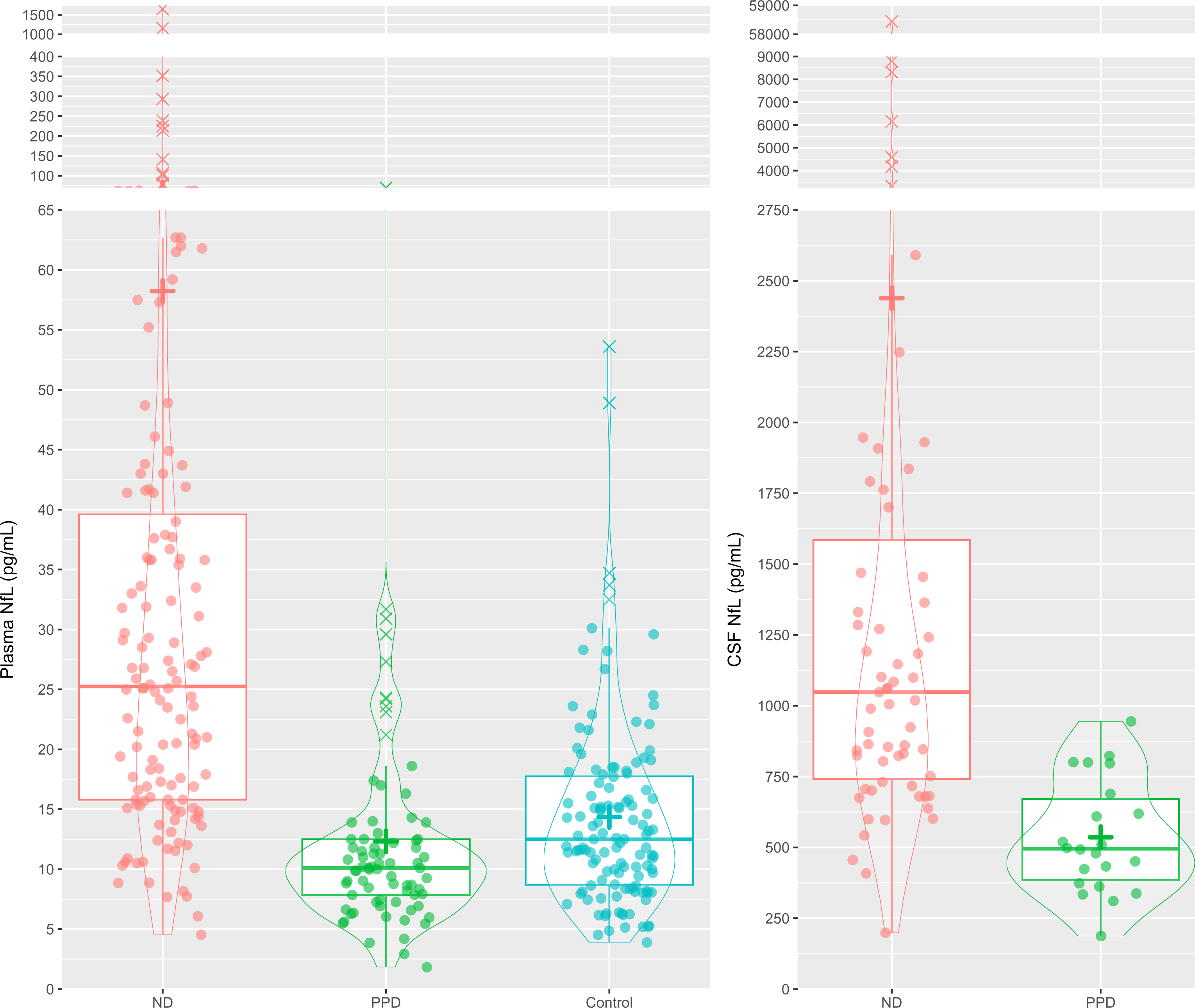
Plasma and CSF neurofilament light chain (NfL) levels in neurodegenerative disorders (ND), primary psychiatric disorders (PPD), and Controls. + = mean level CSF: cerebrospinal fluid; NfL: neurofilament light chain; ND: neurodegenerative disorder; PPD: primary psychiatric disorder

Comparing to controls, plasma NfL levels were higher in ND (median 25.2pg/mL vs 12.5pg/mL, ß: 0.13, 95%CI [0.07, 0.48], p<0.001), but there was no difference between PPD and controls (10.1pg/mL vs 12.5pg/mL, ß: 0.00 [-0.03, 0.03], p=0.998).

ROC curve analyses (Figure 2, Table 2) demonstrated strong diagnostic performance of plasma NfL in distinguishing ND from PPD, with an area under the curve (AUC) of 0.86 [0.81, 0,92], with an optimal cut-off of 14.1pg/mL associated with 81% specificity, 85% sensitivity, 4.34 positive likelihood ratio (LR+), 0.19 negative likelihood ratio (LR-), 88% positive predictive value (PPV), 75% negative predictive value (NPV), **22.6** diagnostic odds ratio (DOR), 83% accuracy (base rate/disease prevalence 63.85%).

**Figure 2.**
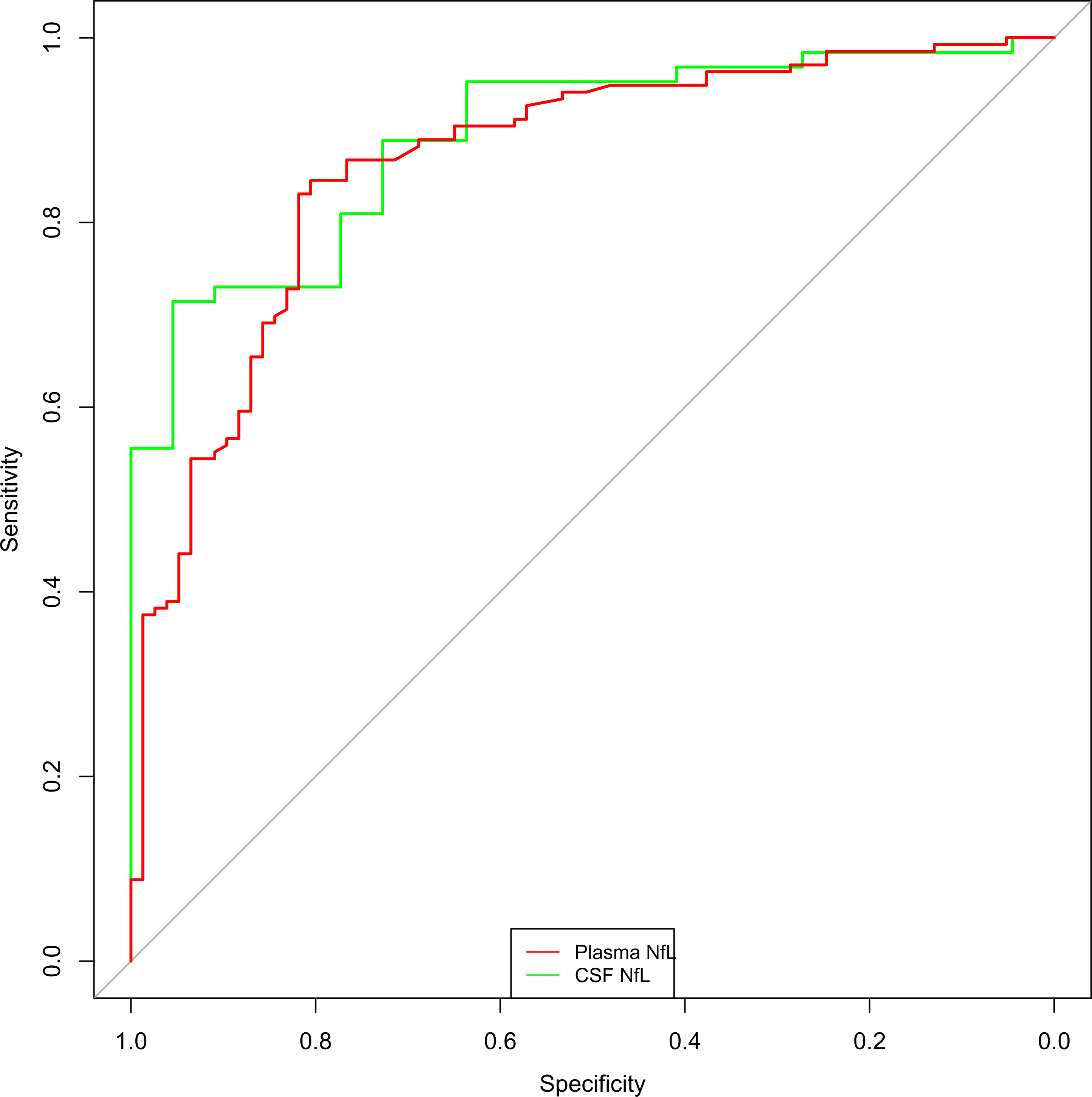
Receiver operator characteristic analysis curves for plasma and CSF neurofilament light (NfL).

**Table 2.**
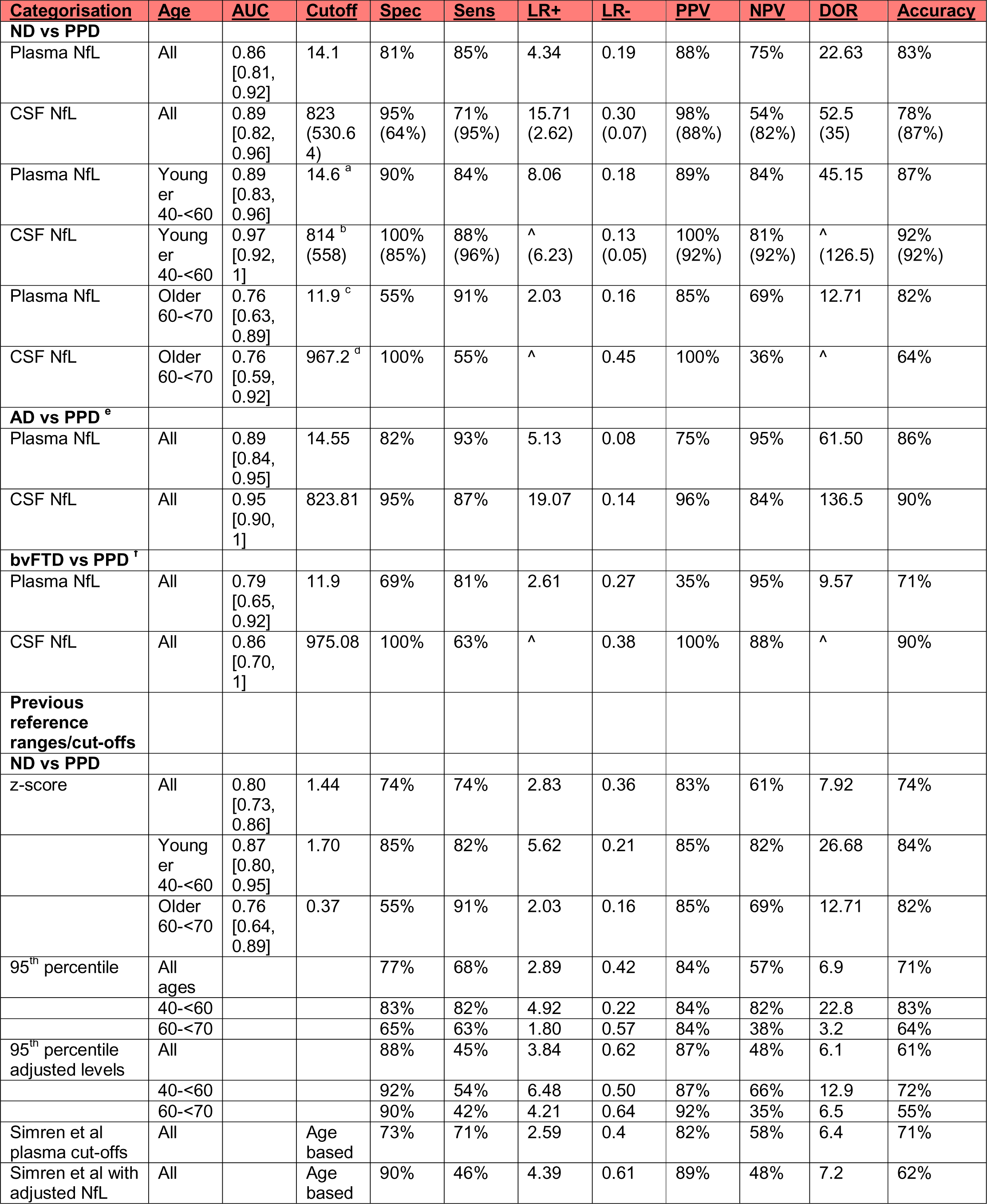
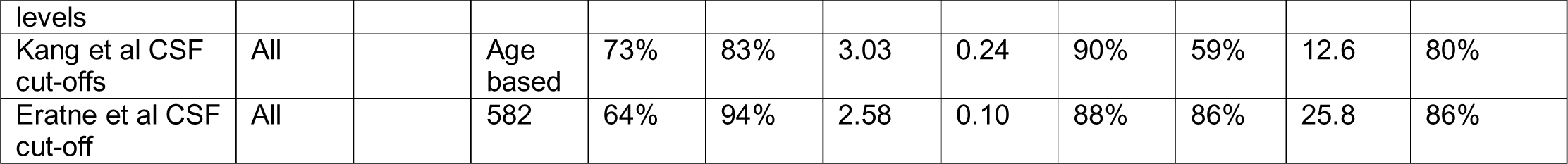
Details of ROC curve analyses and diagnostic test parameters. AD: Alzheimer disease; AUC: area under the curve; bvFTD: behavioural variant frontotemporal dementia; CSF: cerebrospinal fluid; LR+: positive likelihood ratio; LR-: negative likelihood ratio; NfL: neurofilament light chain; ND: neurodegenerative disorder; NPV: negative predictive value; PPD: primary psychiatric disorder; PPV: positive predictive value; Sens: sensitivity; Spec: specificity ^: value of infinity / not able to be calculated (usually because PPV was 100%) a: Alternative cut-offs optimising specificity were: 30.8pg/mL (100% specificity), 24.4pg/mL (98% specificity), 24pg/mL (96% specificity), 14.6pg/mL (90%), and for sensitivity: 6.02pg/mL (100%), 8.04pg/mL (98%), 10.1pg/mL (94%), 11.5pg/mL (90%). b: Alternative CSF cutoffs for specificity were 814pg/mL (100% specificity), 743pg/mL (92%), 558pg/mL (85%), and for sensitivity were 445pg/mL (100% sensitivity), 558pg/mL (96% sensitivity, 85% specificity), 638pg/mL (92%). c: Alternative cut-offs associated with 100%, 95%, 90% specificity were 74.9pg/mL, 31.8pg/mL, 31pg/mL, respectively. Alternative cut-offs optimising for sensitivity were 7.59pg/mL (100% sensitivity), 10.35pg/mL (98%), 10.85pg/mL (95%), 11.9pg/mL (91%). d: Alternative cut-offs optimising for specificity were 967pg/mL (100%) and 823pg/mL (88%), and for sensitivity were 511pg/mL (97%), 571pg/mL (94%), 600pg/mL (90%). e: Higher DOR and accuracy in younger people (154 and infinity, 91% and 96%, for plasma and CSF NfL respectively) f: Higher DOR and accuracy in younger people (16.5 and infinity, 86% and 94%, for plasma and CSF NfL respectively)

CSF NfL had a similar AUC (0.89 [0.82, 0.96]). The optimal CSF cut-off of 823pg/mL was associated with 95% specificity, 71% sensitivity, 15.71 LR+, 0.30 LR-, 98% PPV, 54% NPV, 52.5 DOR, 78% accuracy. An alternative CSF cut-off, optimising for sensitivity was 531pg/mL, 64% specificity, 95% sensitivity, 2.62 LR+, 0.07 LR-, 88% PPV, 82% NPV, 35 DOR, 87% accuracy. There was no statistical difference between plasma NfL and CSF performance (p=0.520).

Considering specific ND subgroups that are the most common differential diagnoses, AD and bvFTD, plasma and CSF NfL had high diagnostic performance for all ages (AD vs PPD AUCs 0.89 (plasma) and 0.95 (CSF); bvFTD vs PPD AUCs 0.79 (plasma), 0.86 (CSF)), with even stronger performance in younger people (full details in Table 2).

### Aim 2: Age-based cut-offs and cut-offs optimised for screening

We investigated cut-offs and diagnostic performance in younger people (40-<60 years), and older people (60-<70 years), similar to recent publications [21]. We restricted to these age ranges as these had the greatest overlap between ND and PPD. In addition, we assessed alternative cutoffs optimised for screening, for 100%, 97.5%, 95%, and 90% specificity, and sensitivity, also similar to recent publications [30]. Alternative cutoffs and additional information are presented in Table 2, Supplementary Material Figures 1-3.

#### Younger people

In younger people (total 98, 48 PPD, 50 ND), plasma NfL had an AUC of 0.89 [0.83, 0.96], cut-off 14.6pg/mL, 90% specificity, 84% sensitivity, 8.06 LR+, 0.18 LR-, 89% PPV, 84% NPV, **45.2 DOR**, 87% accuracy.

CSF NfL had an AUC of 0.97 [0.92, 1.00], 814pg/mL cut-off, 100% specificity, 88% sensitivity, 0.13 LR-, 100% PPV, 81% NPV, 92% accuracy. An alternate cut-off, optimising for sensitivity, was 558pg/mL, 85% specificity, 96% sensitivity, 6.23 LR+, 0.05 LR-, 92% PPV, 92% NPV, 126.5 DOR, 92% accuracy. Plasma and CSF AUCs were not statistically different (p=0.061).

#### Older people

In older people (total 77, 20 PPD, 57 ND), plasma NfL had an AUC of 0.76 [0.63, 0.89], cut-off 11.9pg/mL, 55% specificity, 91% sensitivity, 2.03 LR+, 0.16 LR-, 85% PPV, 69% NPV, **12.7 DOR**, 82% accuracy.

CSF NfL had an AUC of 0.76 [0.59, 0.92], 967pg/mL cut-off, 100% specificity, 55% sensitivity, ∞ LR+, 0.45 LR-, 100% PPV, 36% NPV, 64% accuracy. There were no differences in AUC between plasma and CSF (p=0.961).

Comparing AUCs between younger and older groups, CSF NfL performed better in younger compared to older people (AUC 0.97 vs 0.76, p=0.015), but this was not the case for plasma NfL (0.89 vs 0.76, p=0.081).

### Aim 3: Exploratory comparisons to large reference cohort, different classification systems, and their diagnostic performance

We explored a range of different classification systems, reference ranges and cut-offs, and their diagnostic utility, to understand these important factors and potential issues related to clinical translation.

#### Comparing to large reference control cohort

We compared plasma NfL levels in our cohort to a previously described large reference control cohort (“Control Group 2”) [22,31].

Plasma NfL levels in ND were significantly elevated when compared to Control Group 2, 25.2pg/mL vs 8.34pg/mL, ß: 0.36, 95%CI [0.21, 1.41], p<0.001). Interestingly, levels were also higher in PPD compared to Control Group 2 (PPD ß: 0.04 [0.01, 0.14], p=0.004), and surprisingly, Controls compared to Control Group 2 as well (ß: 0.05 [0.02, 0.18], p<0.001).

This was also reflected in NfL age adjusted z-scores, derived from Control Group 2 as previously described [22]. A large difference was seen between ND and Control Group 2 z-scores (ß: 1.98 [1.81, 2.18], p<0.001). Smaller differences were observed between Control Group 2 and PPD (ß: 0.57 [0.25, 1.07], p<0.001) and Controls (ß: 0.44 [0.29, 0.57], p<0.001).

To investigate the possibility of systematic and analytical bias, factors such as batch effect to explain these surprising findings, especially the differences between Controls and Control Group 2 (i.e. that plasma NfL levels from the batch analysed were systematically higher than the levels from the Control Group 2 batch), we looked at data from 19 samples from our cohort (Batch 1) that were subsequently also analysed on a different batch (Batch 2), using the same analysis kit and platform. Levels between the two batches correlated strongly in a linear fashion (R^2^ = 0.958), but levels in Batch 1 were on average approximately 1.4 times higher than levels in Batch 2.

We then investigated the influence of this possible batch effect by applying a conversion formula and converting NfL levels in the present cohorts (Batch 1), adjusted level=1.33+0.63*x (see Supplementary Table 3 and Supplementary Figure 4). After applying this adjustment, plasma NfL levels were no longer different between PPD and Control Group 2 (ß: -0.02 [-0.06, 0.02], p=0.286), or between the present Controls and Control Group 2 (ß: 0.00 [-0.05, 0.07], p=0.810). Z-scores derived from these adjusted NfL levels were also no longer different between PPD and Control Group 2 (p=0.992), and between the present Controls and Control Group 2 (p=0.176). ND remained elevated however, compared to Control Group 2, even with adjusted levels (17.2pg/mL vs 8.34pg/mL, ß: 0.27 [0.15, 0.79], p<0.001), and the difference in z-scores remained large (ß: 1.44 [1.20, 1.66], p<0.001). This could suggest that a) Batch 2 levels were closer to levels derived from Control Group 2 batches, and b) batch to batch variability could result in potentially spurious findings, where there are small differences between groups.

While the plasma NfL diagnostic performance parameters described above (e.g. AUC, sensitivity, specificity, DOR etc.) were unchanged when using adjusted plasma NfL levels, the values of optimal cut-offs values were influenced. For example, the optimal plasma NfL cutoff for ND vs PPD for all ages changed from 14.1pg/mL to 10.2pg/mL when using the adjusted NfL levels.

For all ages, diagnostic performance of z-scores to distinguish ND from PPD, were outperformed by plasma NfL (AUC 0.80 vs 0.86, p<0.001 and CSF NfL (0.80 vs 0.89, p=0.037). In younger people, z-scores were not statistically different to plasma NfL AUCs (0.87 vs 0.89, p=0.091), but were outperformed by CSF NfL (0.87 vs 0.97, p=0.031). In older people, there were no statistical differences in AUCs between plasma NfL and z-scores (p=0.856), or CSF NfL and z-scores (p=0.948). Using z-scores based on adjusted NfL values did not result in improved AUCs (0.78 for all ages (versus 0.80 unadjusted), 0.87 for younger (vs 0.87), and 0.77 for older (vs 0.77)).

#### Compared to previously described cut-offs

We explored the diagnostic utility of other previously described plasma and CSF cut-offs [3,5,31]. Age-based plasma NfL cut-offs presented by Simrén et al [31] resulted in 73% specificity, 71% sensitivity, 2.59 LR+, 0.40 LR-, 82% PPV, 58% NPV, 6.4 DOR, 71% accuracy. Use of adjusted NfL levels (as describe above) resulted in improved specificity: 90% specificity, 46% sensitivity, 4.39 LR+, 0.61 LR-, 89% PPV, 48% NPV, 7.2 DOR, 62% accuracy.

Using our previously described age-based CSF NfL cut-offs [3] resulted in 73% specificity, 83% sensitivity, 3.03 LR+, 0.24 LR-, 90% PPV, 59% NPV, 12.6 DOR, 80% accuracy. Using only the 582pg/mL cut-off across all ages previously described [5], resulted in 64% specificity, 94% sensitivity, 2.58 LR+, 0.10 LR-, 88% PPV, 78% NPV, 25.8 DOR, 86% accuracy. Once again, this improved in younger people: 85% specificity, 96% sensitivity, 6.23 LR+, 0.05 LR-, 92% PPV, 92% NPV, 126.5 DOR, 92% accuracy.

## DISCUSSION

This study investigated the ability of plasma and CSF NfL to distinguish between diverse ND and PPD in patients referred to and assessed at a specialist clinic. We found significantly elevated plasma and CSF NfL levels in ND compared to PPD, and strong and comparable diagnostic performance of both plasma and CSF NfL to aid in this common, challenging clinical distinction. Diagnostic performance was especially high in younger people, and we described a range of cut-offs based on age and optimising for sensitivity and specificity. Finally, findings from exploratory analyses highlighted potential issues with sample analysis and choices of reference range, issues critical for future broad clincal implementation.

Strengths of this study included focusing on distinguishing diverse ND from a large group of diverse PPD, the clinical and younger nature of cohort. To our knowledge, other studies have not thus far investigated plasma NfL in distinguishing diverse ND from as large a group of well described PPD from a real-world setting. We included a clinical cohort of patients, with no exclusion criteria, to ensure that the findings were reflective of a real-world setting. Findings in diverse conditions and ages in a clinical setting provides important evidence for real-world performance, since current real-world clinical practice involves broad differentials for people with symptoms rather than distinguishing ND from controls or distinguishing only AD from controls or preclinical AD. Understanding NfL levels in diverse PPD from clinical settings is important, since a significant proportion of people who present to clinical services with neuropsychiatric symptoms for assessment of a possible ND (especially younger onset) will be diagnosed with a PPD, and finally, since PPD may be associated with subtle abnormalities and cannot be assumed to be equivalent to or comparable to healthy controls [21,22]. Furthermore, we focused on a relatively younger cohort, in contrast to most previous studies in clinical settings, which have had older cohorts [14,16,20]. This is an important group to investigate; the range of differential diagnoses and atypical presentations means that rates of misdiagnosis and diagnostic uncertainty are all higher in younger people compared to older people [2,32,33].

Diagnostic performance and metrics of both plasma and CSF NfL were very high in younger people, and were higher compared to older people, consistent with previous studies [3,21]. The AUC of CSF NfL in younger people was particularly high (AUC 0.97). This difference was not statistically different from the AUC for plasma NfL in younger people (AUC 0.89), suggesting that plasma NfL levels alone may be sufficient for diagnosis, and there may be little benefit in routinely adding on CSF NfL for a younger person who has already had their plasma NfL levels analysed. Performance was weaker in older people for both plasma and CSF NfL; however, we still found diagnostic utility of a single cut-off for NfL in older people, unlike other studies that did not [21].

Our study results support using plasma NfL as a less invasive alternative to CSF NfL for differentiating ND from PPD, and using age-based cut-offs optimised for sensitivity and sensitivity to aid interpretations. Levels above or below a single optimal cut-off would still offer strong diagnostic utility, especially in younger people; however, greater caution would be required for older people. For example, very high plasma NfL could indicate a neurodegenerative cause (and dismiss a primary psychiatric cause) of a patient’s symptoms and facilitate precision use of more specific investigations based on ND differential diagnoses (such as plasma ptau217 for Alzheimer’s disease [34,35]). Conversely, very low plasma NfL could indicate a primary psychiatric cause of symptoms. Although plasma NfL outperformed CSF NfL in terms of AUC, CSF NfL had very high DOR and accuracy for AD vs PPD and bvFTD vs PPD distinctions. Future research is needed to determine in what circumstances diagnosis would benefit from using both plasma and CSF NfL, such as when ‘borderline’ plasma NfL levels are observed or there remains strong suspicion of ND.

Exploratory comparisons to a large reference cohort revealed some surprising findings, important for future research and clinical implementation. Variability of levels on different batches of resulted in slightly higher levels in all groups in this study, compared to the reference cohort. This did not affect the utility of plasma NfL to distinguish ND from the reference cohort as NfL levels in ND were so highly elevated; however, this further resulted in a possibly spurious finding of elevated levels in PPD (and the present control group) compared to the reference cohort. Adjusting our NfL levels to correct this batch effect reversed this finding of elevated levels in PPD and controls compared to the reference cohort, aligning with previous/expected results [5,22]. In addition, potential systematic batch or analysis factors would influence the actual value of the reference range or cut-off (for example, an optimal cut-off changed from 14.05pg/mL to 10.18pg/mL). These exploratory findings highlight potential limitations and the need for caution in using levels and reference ranges derived from other cohorts and different batches, caution in interpreting individual levels, especially levels relatively close to a reference range/cut-off or ‘borderline’, consistent with other studies that have investigated similar issues [28]. Our findings support the importance of a local control group, contrary to our previous study where comparisions to the large reference cohort performed were comparable and at times were more useful that comparing to a local control group [22]. In addition, the findings of this study would also suggest that raw levels and cut-offs were superior to our previously described age-adjusted z-scores, and age-based cut-offs derived from the large reference cohort [22,31]. Future research should further investigate these potential issues, to advance analysis technologies and improve accurate interpretation of individual levels.

Limitations of this study include the relatively small subset of patients who had both plasma and CSF NfL, and the lack of serial NfL levels. Future studies will include larger numbers of people with both CSF and plasma NfL levels, and investigate diagnostic utility of serial NfL levels. Most patients in this study did not have definitive confirmation, such as genetic or post-mortem confirmation of ND. While patients had comprehensive multidisciplinary assessments and multimodal investigations with current gold standard clinical diagnosis and longitudinal follow up, the limitations and instability of even gold standard clinical diagnosis are recognised [2,3,7]. Of note, CSF NfL in this study’s subset of 85 patients had a slightly lower AUC for all ages compared to our previous study (0.89 vs 0.94), which had a larger sample size and longer follow up duration [5]. The smaller sample sizes in this study could also have contributed to wider confidence intervals and difficulty detecting true differences between AUCs. Therefore, it is possible that with larger sample sizes, additional time and follow up of patients, the diagnostic categorisation and overall findings from this study for both plasma and CSF NfL could be different. The relatively small sample of older people, especially people over age 70, means that our findings in older people must be taken with caution, and replicated in larger studies. Finally, our findings from a specialist service cannot be directly translated to lower prevalence settings such as primary care, and studies are underway in these settings.

To conclude, this study found strong diagnostic performance of both plasma and CSF NfL to distinguish ND from PPD in a real-world clinical setting. NfL had particularly strong diagnostic performance in younger people, where the range of differential diagnoses and atypical presentations, misdiagnosis, and diagnostic delay, are all greater [2,32,33]. The comparability of plasma NfL to CSF NfL adds important evidence for the utility of a simple blood-based biomarker to assist in a common, yet challenging clinical situation, as a screening test for neurodegeneration, aking to a ‘CRP for the brain’, to reduce misdiagnosis and delay and improve precision care and outcomes for patients, their families, and healthcare systems. Future research will need to focus on implementation and translational issues such as analytical, technological, and reference range issues.

## Data Availability

All data produced in the present study are available upon reasonable request to the authors

## ACKNOWLEDGEMENTS AND FUNDING SOURCES

HZ is a Wallenberg Scholar and a Distinguished Professor at the Swedish Research Council supported by grants from the Swedish Research Council (#2023-00356; #2022-01018 and #2019-02397), the European Union’s Horizon Europe research and innovation programme under grant agreement No 101053962, and Swedish State Support for Clinical Research (#ALFGBG-71320).

MK is supported by the Nick Christopher PhD scholarship and the Research Training Program Scholarship from the Department of Psychiatry, University of Melbourne with contributions from the Australian Commonwealth Government, and the Ramsay Hospital Research Foundation.

DE is supported by Mary Stewart Graduate Research Bursary and the Research Training Program Scholarship from the Department of Psychiatry, University of Melbourne with contributions from the Australian Commonwealth Government, and NHMRC Ideas Grant (1185180).

The authors would like to thank all the current and past Neuropsychiatry Centre clinicians. Finally, and most importantly, patients and participants and their families for their participation.

The corresponding author had full access to all the data in the study and had final responsibility for the decision to submit for publication.

## Statistical analysis conducted by

Dr Dhamidhu Eratne

## CONSENT STATEMENT

All human subjects provided informed consent.

## Appendix: collaborators/contributors

### On behalf of others in The MiND Study Group

On behalf of others in The MiND Study Group:
| **Name** | **Affiliations (separated by semi colon)** |
| --- | --- |
| Hannah Dobson | Psychiatry, Alfred Health; Neuropsychiatry, Royal Melbourne Hospital |
| Rosie Watson | Population Health and Immunity Division, The Walter and Eliza Hall Institute of Medical Research, Parkville, Australia;<br>Department of Aged Care and Medicine, The Royal Melbourne Hospital, University of Melbourne, Parkville, Australia |
| Nawaf Yassi | Population Health and Immunity Division, The Walter and Eliza Hall Institute of Medical Research, Parkville, Australia;<br>Department of Medicine and Neurology, Melbourne Brain Centre at the Royal Melbourne Hospital, University of Melbourne, Parkville, Australia |
| Terence J. O'Brien | Department of Neuroscience, The Central Clinical School, Monash University |
| Patrick Kwan | Department of Neuroscience, The Central Clinical School, Monash University |
| Christos Pantelis | Melbourne Neuropsychiatry Centre & Department of Psychiatry, University of Melbourne & Melbourne Health; Mid West Area Mental Health Service, Melbourne Health |
| Oskar Hansson | Department of Clinical Sciences, Clinical Memory Research Unit, Faculty of Medicine, Lund University, Lund/Malmö, Sweden |
| Shorena Janelidze | Department of Clinical Sciences, Clinical Memory Research Unit, Faculty of Medicine, Lund University, Lund/Malmö, Sweden |
| Christopher Fowler | The Florey Institute of Neuroscience and Mental Health, The University of Melbourne, Parkville, VIC |
| Richard Kanaan | Dept of Psychiatry, University of Melbourne, Austin Health, Heidelberg, VIC 3084 |
| Piero Perucca | Department of Medicine, Austin Health, The University of Melbourne; Comprehensive Epilepsy Program, Austin Health; Department of Neuroscience, Central Clinical School, Monash University; Department of Neurology, The Royal Melbourne Hospital; Department of Neurology, Alfred Health, Melbourne, VIC, Australia |
| Anna King | The Wicking Dementia Centre, Tasmania |
| Jane Gunn | Department of General Practice, The University of Melbourne |
| Tianxin Pan | Health Economics Unit \| Centre for Health Policy \| Melbourne School of Population and Global Health, The University of Melbourne |
| Ilias Goranitis | Health Economics Unit \| Centre for Health Policy \| Melbourne School of Population and Global Health, The University of Melbourne |
| Steve Simpson-Yap | CORE, The Royal Melbourne Hospital, The University of Melbourne, Melbourne, VIC, Australia |

## REFERENCES

[1] Loi SM, Goh AMY, Mocellin R, Malpas CB, Parker S, Eratne D, et al. Time to diagnosis in younger-onset dementia and the impact of a specialist diagnostic service. Int Psychogeriatr 2022;34:367–75. 10.1017/S1041610220001489.

[2] Tsoukra P, Velakoulis D, Wibawa P, Malpas CB, Walterfang M, Evans A, et al. The Diagnostic Challenge of Young-Onset Dementia Syndromes and Primary Psychiatric Diseases: Results From a Retrospective 20-Year Cross-Sectional Study. J Neuropsychiatry Clin Neurosci 2022;34:44–52. 10.1176/appi.neuropsych.20100266.

[3] Kang MJY, Eratne D, Dobson H, Malpas CB, Keem M, Lewis C, et al. Cerebrospinal fluid neurofilament light predicts longitudinal diagnostic change in patients with psychiatric and neurodegenerative disorders. Acta Neuropsychiatr 2023:1–12. 10.1017/neu.2023.25.

[4] Eratne D, Loi SM, Walia N, Farrand S, Li Q-X, Varghese S, et al. A pilot study of the utility of cerebrospinal fluid neurofilament light chain in differentiating neurodegenerative from psychiatric disorders: A “C-reactive protein” for psychiatrists and neurologists? Aust N Z J Psychiatry 2020;54:57–67. 10.1177/0004867419857811.

[5] Eratne D, Loi SM, Li Q-X, Stehmann C, Malpas CB, Santillo A, et al. Cerebrospinal fluid neurofilament light chain differentiates primary psychiatric disorders from rapidly progressive, Alzheimer’s disease and frontotemporal disorders in clinical settings. Alzheimer’s & Dementia 2022;18:2218–33. 10.1002/alz.12549.

[6] Eratne D, Loi SM, Li QX, Varghese S, McGlade A, Collins S, et al. Cerebrospinal fluid neurofilament light chain is elevated in Niemann–Pick type C compared to psychiatric disorders and healthy controls and may be a marker of treatment response. Australian and New Zealand Journal of Psychiatry 2020;54:648–9. 10.1177/0004867419893431.

[7] Eratne D, Keem M, Lewis C, Kang M, Walterfang M, Farrand S, et al. Cerebrospinal fluid neurofilament light chain differentiates behavioural variant frontotemporal dementia progressors from non-progressors. Journal of the Neurological Sciences 2022;442:120439. 10.1016/j.jns.2022.120439.

[8] Karantali E, Kazis D, Chatzikonstantinou S, Petridis F, Mavroudis I. The role of neurofilament light chain in frontotemporal dementia: a meta-analysis. Aging Clinical and Experimental Research 2020. 10.1007/s40520-020-01554-8.

[9] Davy V, Dumurgier J, Fayosse A, Paquet C, Cognat E. Neurofilaments as Emerging Biomarkers of Neuroaxonal Damage to Differentiate Behavioral Frontotemporal Dementia from Primary Psychiatric Disorders: A Systematic Review. Diagnostics 2021;11:754. 10.3390/diagnostics11050754.

[10] Forgrave LM, Ma M, Best JR, DeMarco ML. The diagnostic performance of neurofilament light chain in CSF and blood for Alzheimer’s disease, frontotemporal dementia, and amyotrophic lateral sclerosis: A systematic review and metaJanalysis. Alzheimer’s & Dementia: Diagnosis, Assessment & Disease Monitoring 2019;11:730–43. 10.1016/j.dadm.2019.08.009.

[11] Santos F, Cabreira V, Rocha S, Massano J. Blood Biomarkers for the Diagnosis of Neurodegenerative Dementia: A Systematic Review. J Geriatr Psychiatry Neurol 2023;36:267–81. 10.1177/08919887221141651.

[12] Wang S-Y, Chen W, Xu W, Li J-Q, Hou X-H, Ou Y-N, et al. Neurofilament Light Chain in Cerebrospinal Fluid and Blood as a Biomarker for Neurodegenerative Diseases: A Systematic Review and Meta-Analysis. J Alzheimers Dis 2019;72:1353–61. 10.3233/JAD-190615.

[13] Ashton NJ, Janelidze S, Al Khleifat A, Leuzy A, van der Ende EL, Karikari TK, et al. A multicentre validation study of the diagnostic value of plasma neurofilament light. Nat Commun 2021;12:3400. 10.1038/s41467-021-23620-z.

[14] Götze K, Vrillon A, Bouaziz-Amar E, Mouton-Liger F, Hugon J, Martinet M, et al. Plasma neurofilament light chain in memory clinic practice: Evidence from a real-life study. Neurobiology of Disease 2023;176:105937. 10.1016/j.nbd.2022.105937.

[15] Niikado M, Chrem-Méndez P, Itzcovich T, Barbieri-Kennedy M, Calandri I, Martinetto H, et al. Evaluation of Cerebrospinal Fluid Neurofilament Light Chain as a Routine Biomarker in a Memory Clinic. J Gerontol A Biol Sci Med Sci 2019;74:442–5. 10.1093/gerona/gly179.

[16] Sarto J, Ruiz-García R, Guillén N, Ramos-Campoy Ó, Falgàs N, Esteller D, et al. Diagnostic Performance and Clinical Applicability of Blood-Based Biomarkers in a Prospective Memory Clinic Cohort. Neurology 2023;100:e860–73. 10.1212/WNL.0000000000201597.

[17] Verberk IMW, Laarhuis MB, van den Bosch KA, Ebenau JL, van Leeuwenstijn M, Prins ND, et al. Serum markers glial fibrillary acidic protein and neurofilament light for prognosis and monitoring in cognitively normal older people: a prospective memory clinic-based cohort study. The Lancet Healthy Longevity 2021;2:e87–95. 10.1016/S2666-7568(20)30061-1.

[18] Vrillon A, Ashton NJ, Karikari TK, Götze K, Cognat E, Dumurgier J, et al. Comparison of CSF and plasma NfL and pNfH for Alzheimer’s disease diagnosis: a memory clinic study. J Neurol 2024;271:1297–310. 10.1007/s00415-023-12066-6.

[19] Gleerup HS, Simonsen AH, Høgh P. The Added Value of Cerebrospinal Fluid Neurofilament Light Chain to Existing Diagnostic Methods and Biomarkers in a Mixed Memory Clinic Cohort of Consecutive Patients. ADR 2022;6:121–7. 10.3233/ADR-210047.

[20] Willemse EAJ, Scheltens P, Teunissen CE, Vijverberg EGB. A neurologist’s perspective on serum neurofilament light in the memory clinic: a prospective implementation study. Alz Res Therapy 2021;13:101. 10.1186/s13195-021-00841-4.

[21] Light V, Jones SL, Rahme E, Rousseau K, Boer S de, Vermunt L, et al. Clinical Accuracy of Serum Neurofilament Light to Differentiate Frontotemporal Dementia from Primary Psychiatric Disorders is Age-Dependent. The American Journal of Geriatric Psychiatry 2024;0. 10.1016/j.jagp.2024.03.008.

[22] Eratne D, Kang M, Malpas C, Simpson-Yap S, Lewis C, Dang C, et al. Plasma neurofilament light in behavioural variant frontotemporal dementia compared to mood and psychotic disorders. Aust N Z J Psychiatry 2024;58:70–81. 10.1177/00048674231187312.

[23] Katisko K, Cajanus A, Jääskeläinen O, Kontkanen A, Hartikainen P, Korhonen VE, et al. Serum neurofilament light chain is a discriminative biomarker between frontotemporal lobar degeneration and primary psychiatric disorders. Journal of Neurology 2020;267:162–7. 10.1007/s00415-019-09567-8.

[24] Al Shweiki MR, Steinacker P, Oeckl P, Hengerer B, Danek A, Fassbender K, et al. Neurofilament light chain as a blood biomarker to differentiate psychiatric disorders from behavioural variant frontotemporal dementia. Journal of Psychiatric Research 2019;113:137–40. 10.1016/j.jpsychires.2019.03.019.

[25] Arslan B, Zetterberg H. Neurofilament light chain as neuronal injury marker – what is needed to facilitate implementation in clinical laboratory practice? Clinical Chemistry and Laboratory Medicine (CCLM) 2023. 10.1515/cclm-2023-0036.

[26] Coppens S, Lehmann S, Hopley C, Hirtz C. Neurofilament-Light, a Promising Biomarker: Analytical, Metrological and Clinical Challenges. International Journal of Molecular Sciences 2023;24:11624. 10.3390/ijms241411624.

[27] Bavato F, Barro C, Schnider LK, Simrén J, Zetterberg H, Seifritz E, et al. Introducing neurofilament light chain measure in psychiatry: current evidence, opportunities, and pitfalls. Mol Psychiatry 2024:1–17. 10.1038/s41380-024-02524-6.

[28] Sotirchos ES, Hu C, Smith MD, Lord H-N, DuVal AL, Arrambide G, et al. Agreement Between Published Reference Resources for Neurofilament Light Chain Levels in People With Multiple Sclerosis. Neurology 2023. 10.1212/WNL.0000000000207957.

[29] Benkert P, Meier S, Schaedelin S, Manouchehrinia A, Yaldizli Ö, Maceski A, et al. Serum neurofilament light chain for individual prognostication of disease activity in people with multiple sclerosis: a retrospective modelling and validation study. The Lancet Neurology 2022;21:246–57. 10.1016/S1474-4422(22)00009-6.

[30] Brum WS, Cullen NC, Janelidze S, Ashton NJ, Zimmer ER, Therriault J, et al. A two-step workflow based on plasma p-tau217 to screen for amyloid β positivity with further confirmatory testing only in uncertain cases. Nat Aging 2023;3:1079–90. 10.1038/s43587-023-00471-5.

[31] Simrén J, Andreasson U, Gobom J, Suarez Calvet M, Borroni B, Gillberg C, et al. Establishment of reference values for plasma neurofilament light based on healthy individuals aged 5–90 years. Brain Communications 2022;4:fcac174. 10.1093/braincomms/fcac174.

[32] Draper B, Cations M, White F, Trollor J, Loy C, Brodaty H, et al. Time to diagnosis in young-onset dementia and its determinants: the INSPIRED study. International Journal of Geriatric Psychiatry 2016;31:1217–24. 10.1002/gps.4430.

[33] Rossor MN, Fox NC, Mummery CJ, Schott JM, Warren JD. The diagnosis of young-onset dementia. The Lancet Neurology 2010;9:793–806. 10.1016/S1474-4422(10)70159-9.

[34] Eratne D, Li Q-X, Lewis C, Dang C, Kang MJ, Grewal J, et al. Diagnostic utility of plasma ptau217, ptau181, GFAP for Alzheimer disease in a heterogeneous younger onset dementia clinical cohort 2024:2024.04.29.24306586. 10.1101/2024.04.29.24306586.

[35] Ashton NJ, Brum WS, Di Molfetta G, Benedet AL, Arslan B, Jonaitis E, et al. Diagnostic Accuracy of a Plasma Phosphorylated Tau 217 Immunoassay for Alzheimer Disease Pathology. JAMA Neurology 2024. 10.1001/jamaneurol.2023.5319.

