## Supplementary Material for "Plasma and CSF neurofilament light chain distinguish neurodegenerative from primary psychiatric conditions in a clinical setting"

| | ND *N=63* | PPD *N=22* | N |
|---|---|---|---|
| age | 60.9 [57.4;65.0] | 58.6 [51.3;63.7] | 85 |
| sex: Female | 27 (42.9%) | 7 (31.8%) | 85 |
| weight | 74.2 [63.2;91.7] | 82.0 [79.0;86.1] | 67 |
| nfl | 24.4 [15.6;35.8] | 10.3 [7.55;12.3] | 85 |
| lognfl | 1.39 [1.19;1.55] | 1.01 [0.88;1.09] | 85 |
| nfl.z | 2.05 [1.44;2.69] | 0.15 [-0.54;1.25] | 85 |
| nfl.percentile | 0.98 [0.92;1.00] | 0.56 [0.30;0.89] | 85 |
| nfl_csf_pilot2024 | 1048 [741;1585] | 495 [386;671] | 85 |
| logcsfnfl | 3.02 [2.87;3.20] | 2.69 [2.59;2.83] | 85 |

**Supplementary Table 1. Subset with paired CSF and plasma**
Data are median [interquartile range] or n (%).

| | ND *N=73* | PPD *N=55* | Control *N=124* | N |
|---|---|---|---|---|
| age | 60.8 [52.8;67.8] | 54.3 [43.7;59.4] | 63.2 [56.0;70.0] | 252 |
| sex: Female | 32 (43.8%) | 32 (58.2%) | 88 (71.0%) | 252 |
| weight | 75.8 [58.4;86.0] | 87.0 [71.5;100] | 76.0 [66.0;84.0] | 183 |
| nfl | 26.8 [16.6;43.0] | 10.1 [8.05;12.8] | 12.5 [8.70;17.8] | 252 |
| lognfl | 1.43 [1.22;1.63] | 1.00 [0.91;1.11] | 1.10 [0.94;1.25] | 252 |
| nfl.z | 2.36 [1.34;3.12] | 0.70 [-0.01;1.53] | 0.47 [-0.13;1.24] | 252 |
| nfl.percentile | 0.99 [0.91;1.00] | 0.76 [0.50;0.94] | 0.68 [0.45;0.89] | 252 |
| nfl_csf_pilot2024 | . | . | . | 0 |
| logcsfnfl | . | . | . | 0 |

**Supplementary Table 2. Subset with plasma and no CSF**
Data are median [interquartile range] or n (%).

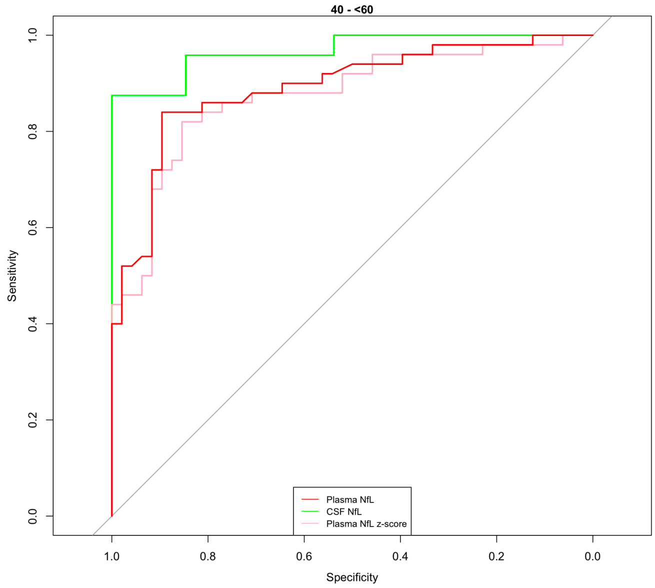


**Supplementary Figure 1. ROC analysis for younger people (40 - <60yo)**

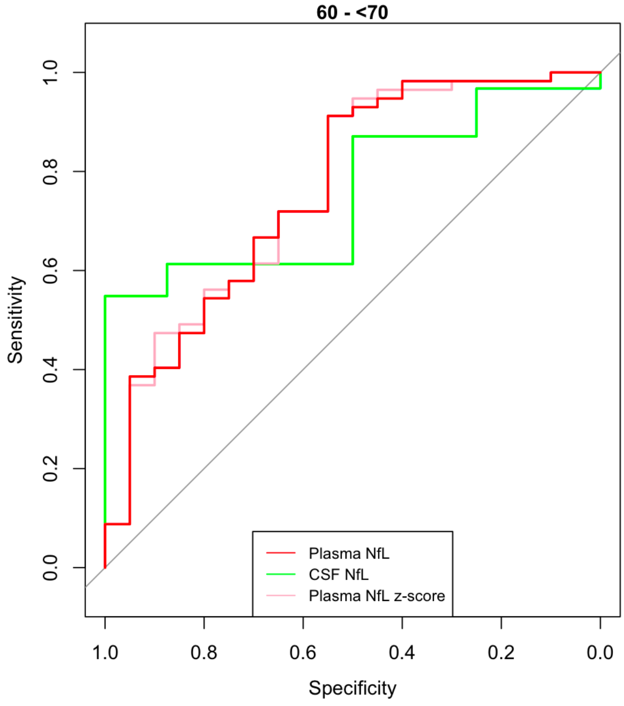


**Supplementary Figure 2. ROC analysis for older people 60 - <70**

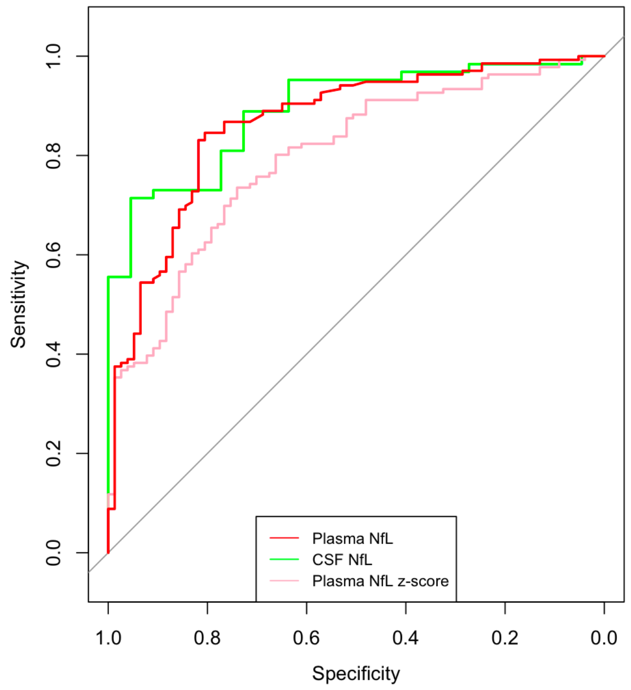


**Supplementary Figure 3. ROC analysis for all ages including z-scores**

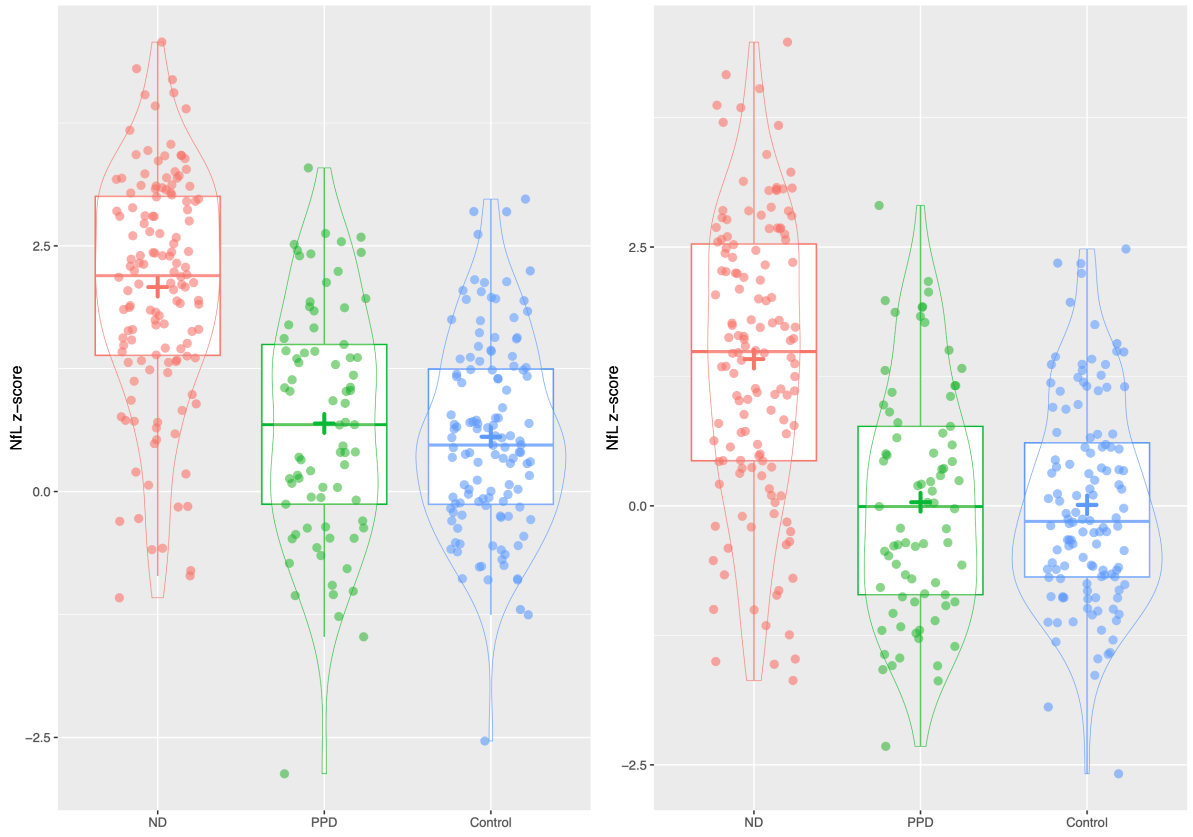


**Supplementary Figure 4. Z-scores before adjustment/conversion (left) and after adjustment/conversion (right).**

| | ND | PPD | Control | Control Group 2 | N |
|---|---|---|---|---|---|
| | *N=136* | *N=77* | *N=124* | *N=1926* | |
| age | 60.8 [55.9;65.9] | 54.8 [46.6;61.8] | 63.2 [56.0;70.0] | 56.0 [48.0;64.0] | 2263 |
| sex: Female | 59 (43.4%) | 39 (50.6%) | 88 (71.0%) | 1218 (63.2%) | 2263 |
| weight | 75.0 [59.8;89.1] | 84.0 [73.4;98.8] | 76.0 [66.0;84.0] | . [.;.] | 250 |
| nfl | 25.2 [15.8;39.6] | 10.1 [7.85;12.5] | 12.5 [8.70;17.8] | 8.34 [6.10;11.6] | 2263 |
| lognfl | 1.40 [1.20;1.60] | 1.00 [0.89;1.10] | 1.10 [0.94;1.25] | 0.92 [0.79;1.06] | 2263 |
| nfl.z | 2.20 [1.39;3.00] | 0.68 [-0.13;1.50] | 0.47 [-0.13;1.24] | -0.01 [-0.70;0.71] | 2263 |
| nfl.percentile | 0.99 [0.92;1.00] | 0.75 [0.45;0.93] | 0.68 [0.45;0.89] | 0.50 [0.24;0.76] | 2263 |
| nfl.adjusted | 17.2 [11.3;26.3] | 7.69 [6.27;9.20] | 9.93 [7.77;14.3] | 8.34 [6.10;11.6] | 2263 |
| lognfl.adjusted | 1.24 [1.05;1.42] | 0.89 [0.80;0.96] | 1.00 [0.89;1.16] | 0.92 [0.79;1.06] | 2263 |
| nfl.adjusted.z | 1.49 [0.44;2.53] | -0.01 [-0.86;0.77] | -0.15 [-0.69;0.61] | -0.01 [-0.70;0.71] | 2263 |
| nfl.adjusted.percentile | 0.93 [0.67;0.99] | 0.50 [0.20;0.78] | 0.44 [0.25;0.73] | 0.50 [0.24;0.76] | 2263 |

**Supplementary Table 3. Adjusted plasma NfL levels and details of Control Group 2**
